# Chronic acid-suppressant use and risk of Oesophageal cancer: protocol for a longitudinal study using a large population based cohort

**DOI:** 10.1101/2021.02.15.21251618

**Authors:** Julia Hippisley-Cox, Xue W Mei, Pui San Tan, Rebecca Fitzgerald, Carol Coupland, Bhagabati Panday-Ghimire, Judith Offman, Peter Sasieni

## Abstract

**Introduction:** Oesophageal cancer is the sixth most common cause for cancer related deaths with over 450,000 new cases and 400,000 resulting deaths per year globally. Most cases in the UK are adenocarcinoma with some of the poorest outcomes from this cancer type in Europe -- mainly due to late diagnosis. The main risk factor for oesophageal adenocarcinoma is chronic reflux disease and due to the high prevalence and non-specific nature of these symptoms most patients are often managed with acid-reflux medications (e.g. Proton Pump Inhibitors (PPIs)) without referral for endoscopy. For those patients that are referred the endoscopy is normal in over 70% of cases, and there is not enough capacity within the NHS for endoscopy especially considering colon cancer screening.

The primary aim of this project is to improve early identification of individuals at risk of oesophageal cancer and reduce over-use of prescription antacids.

**Methods and analysis:** We will conduct a longitudinal cohort study consisted of adults 40 years and over who are free of oesophageal cancer at study entry, using the QResearch database for data gathered between 2000 and 2020. The main exposure is the use of prescription antacids which includes PPI, H2RA, and other aluminium and magnesium containing antacids. The exposure will be categorised based on active ingredients, dose, and duration of use and will be modelled as a time-varying covariate.

**Ethics and dissemination:** Ethical approval for this project was obtained from the QResearch Scientific Committee [Ref: OX39, project title “DELTA - integrated Diagnostic solution for Early detection of Oesophageal cAncer”]. This project has been supported by patient and public involvement panels. We intend to submit the findings for peer-reviewed publication in an academic journal and disseminate them to the public.

**Strength and limitations of this study:**

- This is an open cohort study comprising a nationally representative sample of English population.
- The cohort consists of GP clinic data linked to hospital records, the English national cancer registry and English national death registry.
- This study has access to detailed information on acid-suppressant prescriptions, allowing analysis with consideration of the specific compound, dose, and duration of exposure.
- This study is limited by high rates of missing data for cancer grade and stage, although completeness has improved in recent years, this will be accounted for using appropriate multiple imputation techniques.

## Background

### Epidemiology of oesophageal cancers

Oesophageal cancer is the 8^th^ most common cancer and one of the deadliest cancers in the world^2^. Although oesophageal squamous cell carcinoma (OSCC) is the predominant histological type worldwide, oesophageal adenocarcinoma (OAC) is more common in developed countries such as United States, Australia, United Kingdom, and Western Europe ^2^.

Incidence of the cancer type OAC has increased 6-fold since the 1990s and carries a dismal prognosis. The UK has some of the worst outcomes from this disease in Europe. Clinical guidelines have focussed on minimising endoscopy referrals unless patients have “alarm symptoms” suggestive of cancer. Nevertheless, General Practice referral rates vary widely, and low endoscopy referral rates have been linked with poor outcomes from oesophageal cancer.

A major risk factor for this cancer is chronic heartburn caused by reflux. Three to six percent of individuals with reflux predominant symptoms may have the precursor lesion called Barrett’s oesophagus, but only around 20% of patients with Barrett’s are diagnosed. It is estimated that the burden of OAC could be reduced by up to 50% as a result of increasing the proportion of individuals with reflux symptoms who are investigated. This is a formidable task since heartburn symptoms affect between 5%-20% of the population and account for up to 10% of GP consultations. GPs therefore focus on controlling reflux symptoms with acid-suppressant medication, particularly proton pump inhibitor therapy (PPI). PPIs are highly effective, but patients often continue taking them life-long and there are increasing concerns about long-term side effects including osteoporosis, pneumonia^3 4^ and recently allergy^3^.

### Introduction to DELTA

In 2008 the Chief Medical Officer, Sir Liam Donaldson, raised oesophageal cancer as a public health concern and identified an urgent need to develop a need for a safe, minimally invasive, affordable test applicable to the office setting to diagnose Barrett’s oesophagus. Cambridge University have developed a new minimally invasive test for patients with reflux that can be performed in the GP surgery. This test is called Cytosponge™ - TFF3 which has been tested in over 4,000 individuals across 3 continents.

Our vision is to re-design the clinical pathway for reflux. In primary care a new algorithm applied to NHS prescribing databases will flag symptomatic individuals at risk for oesophageal cancer to their GP. Individuals most at risk will be offered a Cytosponge™ test in a nurse led clinic. The sample will be sent to a centralised laboratory for processing. The H&E and TFF3 stained cells collected by the Cytosponge™ will be assessed using Artificial Intelligence to increase the throughput and reduce the cost. For cases with atypia detected by the pathologist a p53 stain will be added. Individuals at high risk for cancer will be referred for endoscopy and PPI use will be rationalised.

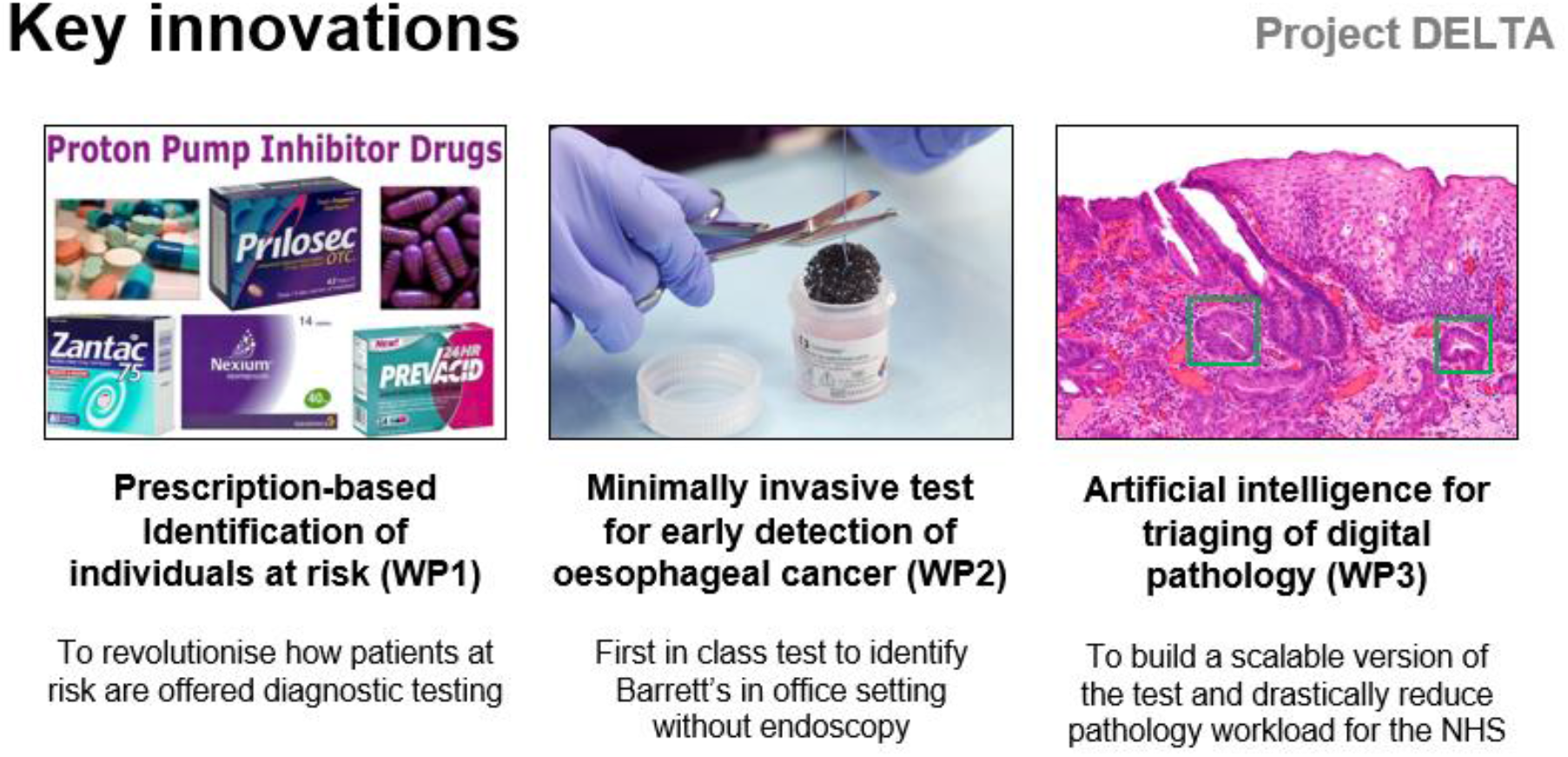

These changes could result in more efficient triage to endoscopy, an economic benefit to the NHS, a social benefit for early detection of a lethal cancer and a reduction in over-use of PPI medication.

### Enhancing the primary care clinical pathway

Currently the clinical pathway for reflux in primary care relies on excluding alarm symptoms (e.g. dysphagia, weight loss) and managing reflux symptoms with acid-suppressant medication. The NICE guidelines recommend endoscopy via the 2 Week Wait pathway if alarm symptoms are present, and routine referral if the symptoms are persistent and occur in the context of other risks factors such as family history (CG27). If a patient is referred to secondary care via the routine pathway the vast majority of patients will be triaged straight to endoscopy. Clinic appointments in secondary care may be led by a dyspepsia nurse or a Gastroenterologist depending on the local policy and resources.

The current, conventional care pathway has a strong focus on triggering endoscopies which results in high costs for the NHS. Additionally, around 70% of these endoscopies are normal with no clinically significant finding (Endoscopy Service Report). The proposed, enhanced care pathway in this project implements a nurse/pharmacist-led clinic for patients with reflux predominant symptoms whereby a Cytosponge procedure is performed in a one-stop visit. This serves as an intermediate step in which the Cytosponge results, together with the high specificity of the test, are used to avoid unnecessary endoscopies.

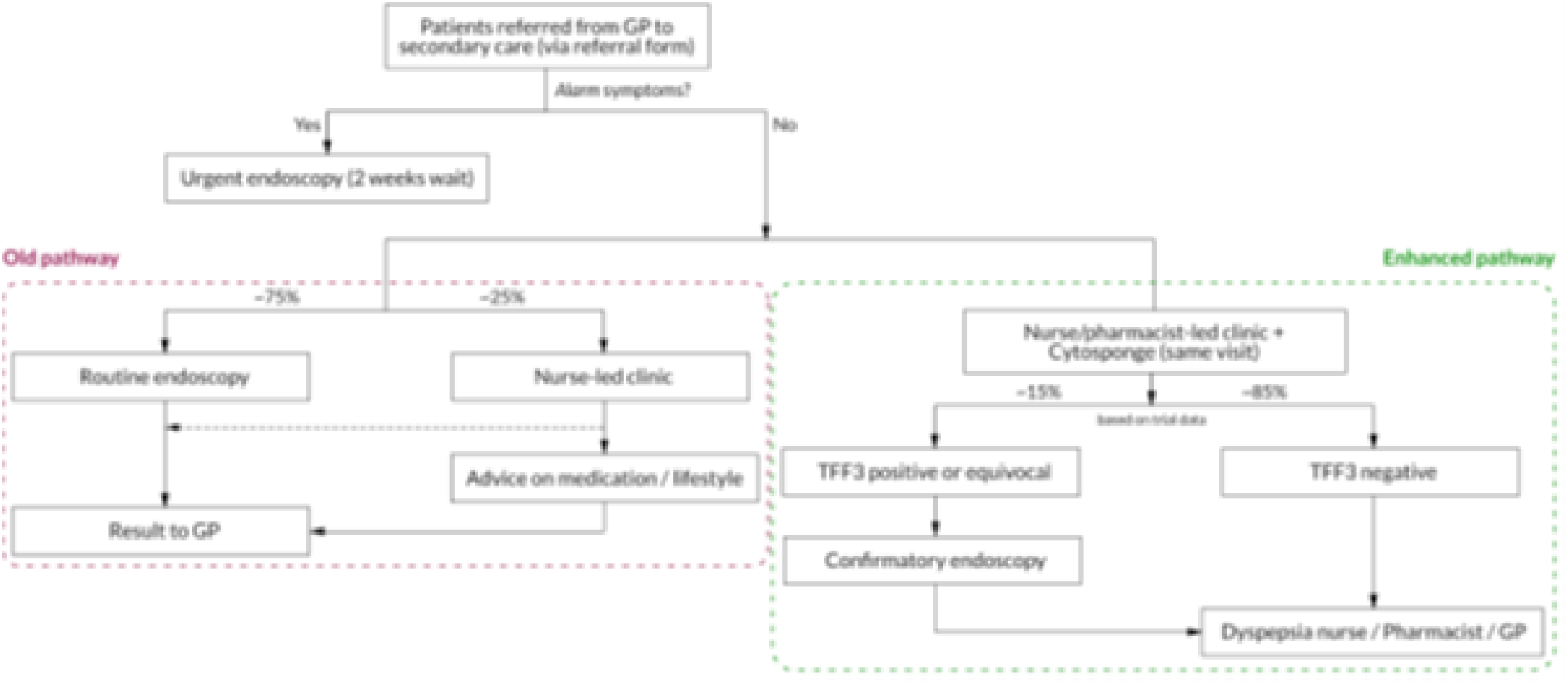

The Cytosponge™ can be readily placed into this pathway (see Fig). We propose that primary care individuals at high risk including those undergoing repeat prescriptions for reflux symptoms are considered for a Cytosponge™ test, with more consideration given to their requirement for long term PPI medication as part of this process. The patients could be flagged electronically to a GP and the test performed by the practice nurse. Alternatively, with the expanding role of the community Pharmacist they would be ideally placed to perform this role. Thus, only patients deemed to be at risk for Barrett’s oesophagus (or another oesophageal condition, or gastric IM) ascertained by the Cytosponge™ test would be referred for endoscopy.

### Introduction to cancer risk prediction tools

Over the last 10 years, Hippisley-Cox et al have developed, validated and implemented a novel set of risk prediction algorithms collectively known as the QCancer algorithms which predict the risk of different types of cancer using readily available information from routinely available electronic health records^5–12^.

The first set of algorithms are designed to improve early diagnosis of an existing cancer on the basis of a combination of symptoms and readily available risk factors in order to identify those patients needing urgent investigation and referral^5^.

The second set of algorithms are designed to estimate longer term cancer risks in order to identify high risk asymptomatic patients with combinations of risk factors who might benefit from systematic screening or interventions to reduce their risk^13^.

This project provides the opportunity to introduce improvements to both sets of QCancer algorithms which are now possible due to the increased size of the QResearch database (which will enable us to distinguish between gastric and oesophageal cancer). It also provides an opportunity to explore risks associated with long-term medication use. For example, we can explore risks associated with proton pump inhibitors which are used to treat chronic reflux symptoms which could also potentially mask development of oesophageal cancer; use of NSAIDs which tend to increase reflux symptoms and use of medications such as aspirin^14 15^ or statins^15^ which may lower risk of oesophageal cancer on their own or in combination with PPI^16^ or cyclo-oxygenase inhibitors^15,17^

## Objectives

The primary objective is to update and validate the QCancer algorithm to identify patients at highest risk of oesophageal cancer who may be suitable for assessment using the Cytosponge device.

Once developed and validated, we envisage that the updated algorithm could be used:

1. Within a consultation between the patient and a clinician with the intention of sharing the information with the patient to assess management options including assessment with Cytosponge
2. To electronically risk-stratify populations by applying the algorithm to all patients to then target and recall patients for a medication review and consideration of Cytosponge sponge assessment based on their levels of risk.
3. To inform mathematical modelling of the potential impact of changing the clinical pathway to include risk assessment +/-Cytosponge.
4. Adapted for use by the general public to improve communication and understanding of risk through implementation into web-based tools
5. Use by researchers to help generate new knowledge or insights.

## Methods

### Study design

We will undertake a cohort study in a large population of primary care patients using the latest version of the QResearch^®^ database (currently version 45).

### Data Sources

QResearch is a high-quality research database established in 2002 which has been used extensively for the development of risk prediction tools which are widely used across the NHS ^18–23^ as well as a wide range of high impact epidemiological research^24–26^. QResearch is a large, representative, validated GP practice research database nationally^27^. The database is linked at an individual patient level to hospital admissions data (including intensive care unit data), cancer registrations and mortality records obtained from the Office for National Statistics. The records are linked using a project-specific pseudonymized NHS number. The recording of NHS numbers is valid and complete for 99.8% of QResearch patients, 99.9% for ONS mortality records and 98% for hospital admissions records^28 29^.

This project will use all four linked data sources (GP, hospital, mortality and cancer registry)

### Study Population

#### Practice inclusion

We will include all practices in England who had been using their EMIS computer system for at least a year. We will randomly allocate three quarters of QResearch practices to the derivation dataset and the remaining quarter to a test (validation) dataset.

#### Patient inclusion

We will identify an open cohort of individuals aged 25-90 years who are registered with practices that have been contributing to QResearch for over 12 months on or after 1^st^ January 2000 (study start date).

#### Patient exclusions

Exclusions will broadly match those for WP2 which aims to implement the risk algorithm for recalling patients for the Cytosponge and hence the following will be excluded.

- Patients with a new onset of alarm symptoms (including haematemesis, unexplained weight loss, dysphagia) in the last 3 months before study entry since these are likely to require urgent referral on the 2-Week Wait pathway for endoscopy.
- Recorded diagnosis of a current or previous oro-pharynx, oesophageal or gastro-oesophageal tumour
- Received prior surgical intervention to the oesophagus
- Recorded oesophageal varices, cirrhosis of the liver

Patients with recorded Barrett’s oesophagus will be included and the presence of Barrett’s oesophagus will be evaluated as a risk factor.

#### Study period

Patients will enter the cohort on the latest of the study start date or the date on which they become 25 or 12 months after registering with their GP practice. Patients will be followed up until they develop oesophageal cancer, die, leave the practice or the study end date or a maximum follow up period of 15 years.

#### Primary Outcome

Our primary outcome of interest is the diagnosis of oesophageal cancer during follow up (all types and subdivided into adenocarcinoma or squamous cell cancer). We will ascertain the diagnoses on the basis of a diagnosis recorded in any of the four linked data sources (1) patients GP record (2) on their linked mortality record (3) hospital record or (4) cancer registry record. We will use the earliest recorded date of oesophageal cancer diagnosis on any of the four data sources as the index date.

#### Secondary outcome

Our secondary outcome is the newly onset diagnosis of Barrett’s Oesophagus.

### Predictor variables

We will examine the following candidate predictor variables based on 12 risk factors already in the QCancer tool^13^ (marked with an asterix) as well as risk factors identified in the literature. The predictor list can be amended by future updates as new knowledge on emerging risk factors becomes available.

#### Demographic variables

1. Age (continuous variable)*.
2. Townsend deprivation score*. This is an area-levelcontinuous score based on the patients’ postcode^30^. Originally developed by Townsend^30^, it includes unemployment (as a percentage of those aged 16 and over who are economically active); non-car ownership (as a percentage of all households); non-home ownership (as a percentage of all households) and household overcrowding. These variables are measured for a given area of approximately 120 households, via the 2011 census, and combined to give a “Townsend score” for that area. A greater Townsend score implies a greater level of deprivation.
3. Ethnicity (9 categories)

#### Concurrent medication (at study entry)

1. Proton pump inhibitors (which are used to treat reflux symptoms)
2. H2 blockers (which are used to treat reflux symptoms)
3. NSAIDs (which can worsen reflux symptoms)
4. Aspirin (which may lower OAC risk)^14 16^
5. Statins (which may lower OAC risk)^17 31^
6. COX inhibitors (which may lower OAC risk)^17^
7. Metformin (which may lower OAC risk)^15^
8. HRT (in female only, may lower OAC risk)^15^
9. Other diabetic drugs

#### Lifestyle and family history

1. Smoking status - non-smoker, ex-smoker, light smoker (1-9/day), moderate(10-19/day) or heavy(20+/day)*.
2. Body mass index (continuous variable, z-scores will be used)*.
3. Alcohol use-non-drinker; light drinker (<1 unit/day); moderate (3-6 units/day); heavy (6+ units/day)*.
4. Family history of bowel or gastric or colorectal cancer.

#### Co-morbidities and investigations

1. Gastric cancer
2. Barrett’s oesophagus*
3. Peptic ulcer disease*
4. Gastro-oesophageal reflux disease (including heart burn)
5. Type 1 and type 2 diabetes*
6. Previous blood cancer*
7. Previous breast cancer*
8. Previous oral cancer*
9. Previous pancreatic cancer*
10. Current H.Pylori infection (which may lower OAC risk)
11. Pernicious anaemia
12. Hiatus hernia
13. Anaemia (including Haemoglobin values)
14. Full blood count
15. CT scan abdomen within the previous 5 years
16. CT scan pelvis within the previous 5 years
17. Barium meal/swallow within the previous 5 years
18. Endoscopy within the previous 5 years

All predictor variables will be based on the latest coded information recorded in the GP record prior to entry to the cohort.

### Descriptive analysis

We will produce the following descriptive analyses for comparison with the literature and national statistics from the [CRUK website](https://www.cancerresearchuk.org/health-professional/cancer-statistics/statistics-by-cancer-type/oesophageal-cancer/incidence#heading-Two).

- Crude incidence of oesophageal cancer by age, sex, ethnicity, deprivation and calendar time.
- Age standardised incidence of oesophageal cancer overall and by type.
- Characteristics of cases diagnosed with oesophageal cancer - age at diagnosis, stage, grade, histology, treatment (surgery, radiotherapy, chemotherapy, other).

34. Development of the models

35. Model development overview

We will use the following steps:

1. Development of prognostic models for each outcome within the derivation data.
2. Evaluation of predictive performance in the validation data.

Separate models will be developed and evaluated for males and females.

### Development of the models using the derivation data

For all analyses, the time origin is entry to the study cohort and the risk period of interest is from the time origin up to the first date of diagnosis of oesophageal cancer. We will develop and evaluate the risk prediction equations using established methods^19 32–35^ We will use second degree fractional polynomials (i.e. with up to two powers)^**31**^ to model non-linear relationships for continuous variables (age, body mass index and Townsend score). Models will include interactions between age and predictor variables focussing on predictor variables which apply across the age range where numbers allow.

### Handling of missing data

For all predictor variables, we will use the most recently available value at the time origin. For indicators of co-morbidities and medication use, the absence of information being recorded is assumed to mean absence of the factor in question. There may be missing data in some variables due to never being recorded: ethnicity, Townsend score, body mass index, smoking status and alcohol intake. We will use multiple imputation with chained equations to replace missing values for these variables ^36–39^.

Prior to the imputation, a complete-case analysis will be fitted using a model containing only the continuous covariates within the derivation data to derive the fractional polynomial order and corresponding powers. Then a multiple imputation model using chained equations will be fitted in the derivation data and will include all predictor variables along with age interaction terms, the Nelson–Aalen estimators of the baseline cumulative hazard, and the outcome indicators (namely, oesophageal cancer). Separate imputation models will be fitted for men and women. We will carry out 5 imputations as this has a relatively high efficiency^29^ and is a pragmatic approach accounting for the size of the datasets and capacity of the available servers and software.

Each analysis model will be fitted in each imputed data set. We will use Rubin’s rules to combine the model parameter estimates across the imputed datasets^40^.

### Variable selection

We will fit models that include all predictor variables initially and retain variables if they have a hazard ratio of < 0.90 or > 1.10 (for binary variables) and are statistically significant at the 0.01 level. For previous diagnoses of other cancers, we will retain variables which were significant at the 0.05 level since some of the cancers are rare. In order to simplify the models, we will focus on variables for the most common conditions and medications and combine similar variables with comparable hazard ratios where possible. If some predictor variables result in very sparse cells (i.e. with not enough participants or events to obtain point estimates and standard errors), we will combine some of these if clinically similar in nature.

For PPI and H2 blockers medication usage, we will determine the association between risk of oesophageal cancer and type of medication (omeprazole etc), dose and duration of exposure (e.g. < 6 months; 6-11 months; 12-23 months; 24-47 months; 48 months or more). We will examine both usage at baseline and also as a time varying exposure during study follow-up.

### Risk equations

We will use the regression coefficients for each variable from the final model as weights which we will combine with the baseline survivor function evaluated for each yearup to 10 years to derive risk equations over a period of 10 years of follow-up^41^. This will enable us to derive risk estimates for each year of follow-up, with a specific focus on 10-year risk estimates. We will estimate the baseline survivor function based on zero values of centred continuous variables, with all binary predictor values set to zero.

### Validation of the models

In the validation data, we will fit an imputation model to enable imputation of missing values for ethnicity, body mass index, alcohol and smoking status. We will carry out 5 imputations. We will apply the risk equations for males and females obtained from the derivation datato of medication (omeprazole etc), dose and the validation dataand calculate measures of performance.

As in previous studies^42^, we will calculate R^2^ values (explained variation where higher values indicate a greater proportion of variation in survival time explained by the model ^43^), D statistics^44^ (a measure of discrimination which quantifies the separation in survival between patients with different levels of predicted risk where higher values indicate better discrimination), Brier scores, and Harrell’s C statistics at 1, 2, 5 and 10 years and combine these across datasets using Rubin’s rules. Harrell’s C statistic^45^ is a measure of discrimination (separation) which quantifies the extent to which those with earlier events have higher risk scores. Higher values of Harrell’s C indicate better performance of the model for predicting the relevant outcome. A value of 1 indicates that the model has perfect discrimination. A value of 0.5 indicates that the model discrimination is no better than chance.

We will calculate 95% confidence intervals for the performance statistics to allow comparisons with alternative models for the same outcome and across different subgroups.^46^

We will assess calibration of the risk scores by comparing the mean predicted risks with the observed risks by tenth of predicted risk. The observed risks will be obtained using Kaplan-Meier estimates evaluated at 10 years, obtained for men and women.

We will also evaluate these performance measures in 6 pre-specified age groups (25-49; 50-59; 60-69; 70-79; 80+), and in people on long term PPIs.

We will also compare the risk prediction model with a simple algorithm based only on criteria such as age and number of electronic prescriptions of PPI/H2 blockers and consider the use of decision curve analysies.

### Updating of the model using new data

We will update the models to ensure the model remains up to date. The baseline survivor function may change after the widespread introduction of the Cytosponge (for example), so will be updated in future models where possible^32^. Even though it may not be possible to fully account for changes in baseline survival over time the risk scores will give a rank ordering of patients that can be used for risk stratification/identification of high-risk groups.

### Development of risk categories

Since there is no currently accepted threshold for classifying high risk of oesophageal cancer, we will examine the distribution of predicted risks and calculate a series of centile values. For each centile threshold, we will calculate the sensitivity of the risk scores.

### Sample size

Sample size calculations for a risk prediction model aim to ensure precise estimation of model parameters whilst minimising potential overfitting. We have used the criteria of Riley et al.^47^ o derive a minimum sample size of 312,616 men corresponding to 543,952 person-years of follow-up. The number of outcome events needed are 349 assuming up to 70 predictors, an event rate of 0.00064, mean follow up of 1.74 years; timepoint 10 years, a R^2^ value of 0.002013. Similarly, a minimum sample size of 665,050 women corresponding to 1,157,1871 person-years of follow-up. The number of outcome events needed are 359 assuming up to 70 predictors, an event rate of 0.00031; mean follow up of 1.74 years; a R^2^ value of 0.0009468. Hence a minimum sample of just over 1 million men and women would be needed in the derivation dataset.

Collins, et al^48^ suggests externally validating a prognostic model requires at least 100 events and ideally, at least 200.

With over 35 million patients and at least 18,000 incident cases of oesophageal cancer on QResearch, we can confirm we have more than ample data both in the training and validation datasets. We will use all the relevant patients on the database to maximise the power and generalisability of the results. We will use STATA (version 16) for analyses. We will adhere to the TRIPOD statement for reporting^49^.

### Public and patient involvement

Patients will be involved in setting the research question, the outcome measures, the design, implementation and dissemination of the study findings. Patient representatives will also advise on dissemination including the use of culturally appropriate lay summaries describing the research and its results.

### Methodological considerations

#### Strengths

The methods to derive and validate these models are broadly the same as for a range of other widely used clinical risk prediction tools derived from the QResearch database ^18–22^. The strengths and limitations of the approach have already been discussed in detail ^19 22 32 33 50,51^. Key strengths include size, wealth of data on risk factors, good ascertainment of outcomes through multiple record linkage, prospective recording of outcomes, use of an established validated database which has been used to develop many risk prediction tools, and lack of selection, recall and respondent bias and robust analysis. UK general practices have good levels of accuracy and completeness in recording clinical diagnoses and prescribed medications ^52^. We think our study has good face validity since it will be conducted in the setting where most patients in the UK are assessed, treated and followed up.

## Limitations

Limitations of our study include the lack of formal adjudication of diagnoses, potential for misclassification of outcomes depending on testing, information bias, and potential for bias due to missing data. However, our database has linked cancer registry, death registry mortality and hospital admissions data and is therefore likely to have picked up the great majority of cases and death thereby minimising ascertainment bias.

The initial evaluation will be done on a separate set of practices and individuals to those which were used to develop the score although the practices all use the same GP clinical computer system (EMIS – the computer system used by 55% of UK GPs). An independent evaluation willbe a more stringent test and should be done (e.g. using data from different clinical systems or the other countries within the UK), but when such independent studies have examined other risk equations, ^50 51 53 54^ they have demonstrated similar performance compared with the validation in the QResearch database^18 19 32^. Whilst our study population is from England and is representative of the English population, it will need to be locally evaluated if used outside of England.

## Data Availability

Data is available and can be accessed by members of the project team undertaking statistical analysis

## References

1. Fitzgerald RC, di Pietro M, O’Donovan M, et al. Cytosponge-trefoil factor 3 versus usual care to identify Barrett’s oesophagus in a primary care setting: a multicentre, pragmatic, randomised controlled trial. The Lancet 2020;396(10247):333-44. doi:https://doi.org/10.1016/S0140-6736(20)31099-0

2. Pennathur A, Gibson MK, Jobe BA, et al. Oesophageal carcinoma. Lancet 2013;381(9864):400–12. doi:10.1016/s0140-6736(12)60643-6 [published Online First: 2013/02/05]

3. Kinoshita Y, Ishimura N, Ishihara S. Advantages and Disadvantages of Long-term Proton Pump Inhibitor Use. J Neurogastroenterol Motil 2018;24(2):182–96. doi:10.5056/jnm18001

4. Lambert AA, Lam JO, Paik JJ, et al. Risk of community-acquired pneumonia with outpatient proton-pump inhibitor therapy: a systematic review and meta-analysis. PLoS One 2015;10(6):e0128004.

5. Hippisley-Cox J, Coupland C. Identifying patients with suspected gastro-oesophageal cancer in primary care: derivation and validation of an algorithm. British Journalof General Practice 2011;61(592):e707–14. doi:10.3399/bjgp11X606609 [published Online First: 2011/11/08]

6. Hippisley-Cox J, Coupland C. Identifying patients with suspected lung cancer in primary care: derivation and validation of an algorithm. The British journal of generalpractice : the journalof the Royal College of General Practitioners 2011;61(592):e715–23. doi:10.3399/bjgp11X606627 [published Online First: 2011/11/08]

7. Hippisley-Cox J, Coupland C. Identifying women with suspected ovarian cancer in primary care: derivation and validation of algorithm. BMJ 2012;344 doi:10.1136/bmj.d8009

8. Hippisley-Cox J, Coupland C. Identifying patients with suspected colorectal cancer in primary care: derivation and validation of an algorithm. British Journalof General Practice 2012;62(594):e29–e37. doi:10.3399/bjgp12X616346

9. Hippisley-Cox J, Coupland C. Identifying patients with suspected pancreatic cancer in primary care: derivation and validation of an algorithm. The British journal of general practice : the journal of the Royal College of General Practitioners 2012;62(594):e38–e45. doi:10.3399/bjgp12X616355

10. Hippisley-Cox J, Coupland C. Identifying patients with suspected renal tract cancer in primary care: derivation and validation of an algorithm. Br J Gen Pract 2012;62(597):e251–60. doi:10.3399/bjgp12X636074

11. Hippisley-Cox J, Coupland C. Symptoms and risk factors to identify women with suspected cancer in primary care: derivation and validation of an algorithm. Br J Gen Pract 2013;63(606):11–21. doi:10.3399/bjgp13X660733

12. Hippisley-Cox J, Coupland C. Symptoms and risk factors to identify men with suspected cancer in primary care: derivation and validation of an algorithm. Br J Gen Pract 2013;63(606):1–10. doi:10.3399/bjgp13X660724

13. Hippisley-Cox J, Coupland C. Development and validation of risk prediction algorithms to estimate future risk of common cancers in men and women: prospective cohort study. BMJ Open 2015;5(3):e007825. doi:10.1136/bmjopen-2015-007825

14. Qiao Y, Yang T, Gan Y, et al. Associations between aspirin use and the risk of cancers: a meta-analysis of observational studies. BMC Cancer 2018;18(1):288. doi:10.1186/s12885-018-4156-5 [published Online First: 2018/03/15]

15. Snider EJ, Kaz AM, Inadomi JM, et al. Chemoprevention of esophageal adenocarcinoma. Gastroenterol Rep (Oxf) 2020;8(4):253–60. doi:10.1093/gastro/goaa040

16. Jankowski JAZ, de Caestecker J, Love SB, et al. Esomeprazole and aspirin in Barrett’s oesophagus (AspECT): a randomised factorial trial. Lancet 2018;392(10145):400–08.

17. Beales IL, Hensley A, Loke Y. Reduced esophageal cancer incidence in statin users, particularly with cyclo-oxygenase inhibition. World J Gastrointest Pharmacol Ther 2013;4(3):69–79. doi:10.4292/wjgpt.v4.i3.69 [published Online First: 2013/08/07]

18. Hippisley-Cox J, Coupland C, Vinogradova Y, et al. Predicting cardiovascular risk in England and Wales: prospective derivation and validation of QRISK2. BMJ 2008:bmj.39609.449676.25. doi:10.1136/bmj.39609.449676.25

19. Hippisley-Cox J, Coupland C, Robson J, et al. Predicting risk of type 2 diabetes in England and Wales: prospective derivation and validation of QDScore. BMJ 2009;338:b880.. doi:10.1136/bmj.b880

20. Hippisley-Cox J, Coupland C. Derivation and validation of updated QFracture algorithm to predict risk of osteoporotic fracture in primary care in the United Kingdom: prospective open cohort study. BMJ 2012;344(may 22 1):e3427–e27. doi:10.1136/bmj.e3427

21. Hippisley-Cox J, Coupland C. Predicting the risk of Chronic Kidney Disease in Men and Women i n England and Wales: prospective derivation and external validation of the QKidney(R) Scores. BMC Family Practice 2010;11:49.

22. Hippisley-Cox J, Coupland C. Development and validation of risk prediction algorithm (QThrombosis) to estimate future risk of venous thromboembolism: prospective cohort study. BMJ 2011;343:d4656. doi:10.1136/bmj.d4656 [published Online First: 2011/08/19]

23. Clift AK, Coupland CAC, Keogh RH, et al. Living risk prediction algorithm (QCOVID) for risk of hospital admission and mortality from coronavirus 19 in adults: national derivation and validation cohort study. BMJ 2020;371:m3731. doi:10.1136/bmj.m3731 [published Online First: 2020/10/22]

24. Coupland CAC, Hill T, Dening T, et al. Anticholinergic Drug Exposure and the Risk of Dementia: A Nested Case-Control Study. JAMA Intern Med 2019 doi:10.1001/jamainternmed.2019.0677 [published Online First: 2019/06/25]

25. Vinogradova Y, Coupland C, Hippisley-Cox J. Use of hormone replacement therapy and risk of venous thromboembolism: nested case-control studies using the QResearch and CPRD databases. BMJ 2019;364:k4810. doi:10.1136/bmj.k4810 [published Online First: 2019/01/11]

26. Vinogradova Y, Coupland C, Hippisley-Cox J. Use of combined oral contraceptives and risk of venous thromboembolism: nested case-control studies using the QResearch and CPRD databases. BMJ 2015;350:h2135. doi:10.1136/bmj.h2135

27. Kontopantelis E, Stevens RJ, Helms PJ, et al. Spatial distribution of clinical computer systems in primary care in England in 2016 and implications for primary care electronic medical record databases: a cross-sectional population study. BMJ Open 2018;8(2) doi:10.1136/bmjopen-2017-020738

28. Hippisley-Cox J, Coupland C. Predicting risk of emergency admission to hospital using primary care data: derivation and validation of QAdmissions score. BMJ Open 2013;3(8):e003482. doi:10.1136/bmjopen-2013-003482 [published Online First: 2013/08/21]

29. Hippisley-Cox J. Validity and completeness of the NHS Number in primary and secondary care electronic data in England 1991-2013. 2013; 1. Hippisley-Cox J. Validity and completeness of the NHS number in primary and secondary care: electronic data in England 1991-2013 http://eprints.nottingham.ac.uk/3153/1/Validity%26CompletenessNHSNumber.pdf (accessed June 2013).

30. Townsend P, Davidson N. The Black report. London: Penguin 1982.

31. Thomas T, Loke Y, Beales ILP. Systematic Review and Meta-analysis: Use of Statins Is Associated with a Reduced Incidence of Oesophageal Adenocarcinoma. J Gastrointest Cancer 2018;49(4):442–54. doi:10.1007/s12029-017-9983-0

32. Hippisley-Cox J, Coupland C. Predicting risk of osteoporotic fracture in men and women in England and Wales: prospective derivation and validation of QFractureScores. BMJ 2009;339:b4229. doi:10.1136/bmj.b4229

33. Hippisley-Cox J, Coupland C, Vinogradova Y, et al. Performance of the QRISK cardiovascular risk prediction algorithm in an independent UK sample of patients from general practice: a validation study. Heart 2008;94:34–39. doi:10.1136/hrt.2007.134890

34. Hippisley-Cox J, Coupland C, Brindle P. Development and validation of QRISK3 risk prediction algorithms to estimate future risk of cardiovascular disease: prospective cohort study. BMJ 2017;357:j2099. doi:10.1136/bmj.j2099

35. Hippisley-Cox J, Coupland C. Development and validation of QDiabetes-2018 risk prediction algorithm to estimate future risk of type 2 diabetes: cohort study. BMJ 2017;359:j5019. doi:10.1136/bmj.j5019

36. Schafer J, Graham J. Missing data: our view of the state of the art. Psychological Methods 2002;7:147–77.

37. Group TAM. Academic Medicine: problems and solutions. BMJ 1989;298:573–79.

38. Steyerberg EW, van Veen M. Imputation is beneficial for handling missing data in predictive models. J Epidemiol Community Health 2007;60:979.

39. Moons KGM, Donders RART, Stijnen T, et al. Using the outcome for imputation of missing predictor values was preferred. J Epidemiol Community Health 2006;59:1092.

40. Rubin DB. Multiple Imputation for Non-response in Surveys. New York: John Wiley 1987.

41. Hosmer D, Lemeshow S. Applied Logistic Regressopm. New York: John Wiley & Sons, Inc. 1989.

42. Hippisley-Cox J, Coupland C, Brindle P. The performance of seven QPrediction risk scores in an independent external sample of patients from general practice: a validation study. BMJ Open 2014;4(8):e005809. doi:10.1136/bmjopen-2014-005809

43. Royston P. Explained variation for survival models. Stata J 2006;6:1–14.

44. Royston P, Sauerbrei W. A new measure of prognostic separation in survival data. Stat Med 2004;23:723–48.

45. Harrell F, Lee K, Mark D. Multivariable prognostic models: issues in developing models, evaluating assumptions and adequacy, and measuring and reducing errors. Stat Med 1996;15:361 –87.

46. Newson RB. Comparing the predictive powers of survival models using Harrell’s C or Somers’ D. Stata Journal 2010;10(3):339–58.

47. Riley RD, Ensor J, Snell KIE, et al. Calculating the sample size required for developing a clinical prediction model. BMJ 2020;368:m441. doi:10.1136/bmj.m441 [published Online First: 2020/03/20]

48. Collins GS, Ogundimu EO, Altman DG. Sample size considerations for the external validation of a multivariable prognostic model: a resampling study. Stat Med 2015 doi:10.1002/sim.6787 [published Online First: 2015/11/11]

49. Collins GS, Reitsma JB, Altman DG, et al. Transparent Reporting of a multivariable prediction model for Individual Prognosis Or Diagnosis (TRIPOD): The TRIPOD StatementThe TRIPOD Statement. Annals of Internal Medicine 2015;162(1):55–63. doi:10.7326/M14-0697

50. Collins GS, Mallett S, Altman DG. Predicting risk of osteoporotic and hip fracture in the United Kingdom: prospective independent and external validation of QFractureScores. BMJ 2011;342:d3651.

51. Collins GS, Altman DG. External validation of the QDScore for predicting the 10-year risk of developing Type?2 diabetes. Diabetic Medicine 2011;28:599–607. doi:10.1111/j.1464-5491.2011.03237.x

52. Majeed A. Sources, uses, strengths and limitations of data collected in primary care in England. Health stat 2004(21):5–14.

53. Collins GS, Altman DG. Predicting the 10 year risk of cardiovascular disease in the United Kingdom: independent and external validation of an updated version ofQRISK2. BMJ 2012;344:e4181. doi:10.1136/bmj.e4181

54. Collins GS, Altman DG. An independent and external validation of QRISK2 cardiovascular disease risk score: a prospective open cohort study. BMJ 2010;340:c2442. doi:10.1136/bmj.c2442

